# Behavioral and Healthcare Determinants of Self-Reported Scabies in Chiwanda Ward, Nyasa District, Tanzania

**DOI:** 10.64898/2026.02.25.26347071

**Authors:** I Kilagwa, N Renfrid, M Mwabukusi, Hafidh S. Hassan, Mwingira Victor, Athumani M. Lupindu, Sharadhuli. I Kimera

**Author notes:** The material contained in this chapter has been submitted and is under review by PLOS Neglected Tropical Diseases.

## Abstract

**Introduction:** Scabies, caused by Sarcoptes scabiei, is a neglected tropical disease that disproportionately affects underserved rural communities, where transmission is commonly sustained through prolonged close contact and sharing of personal items. This study assessed household scabies experience and associated factors during a past outbreak in Nyasa District.

**Methods:** A retrospective community-based cross-sectional study was conducted among 198 households from four villages. Data were collected using an AfyaData-digitized, expert-validated questionnaire aligned with the International Alliance for the Control of Scabies criteria to improve syndromic specificity. Quantitative data were analyzed using univariable and multivariable logistic regression. Open-ended responses (Q24–Q26) were analyzed thematically and triangulated with regression findings.

**Results:** Overall, 60.6% (120/198) of households reported scabies experience during the outbreak period. In multivariable analysis, higher odds of household scabies experience were associated with sharing personal items (Rarely AOR = 4.059; Frequent AOR = 4.688) and receiving treatment during the outbreak (AOR = 4.705). Non-collaboration with healthcare personnel showed increased odds but was not statistically significant (AOR = 2.035; p = 0.098). Lower odds were observed among households reporting “not sure” responses for prior scabies history (AOR = 0.235) and treatment (AOR = 0.249), suggesting uncertainty-related misclassification. Qualitative themes mapped to these determinants: sharing items aligned with laundry/bedding and clothing hygiene practices (“…kufua… na kuzidisha usafi”); treatment aligned with effectiveness concerns and access/availability barriers (“madawa… hayatibu”; “matibabu mbali”); collaboration aligned with requests for outreach/education and follow-up (“watoa huduma hawakufika…”); and “not sure” responses aligned with misconceptions and uncertainty (“tulifikiri… tumerogwa”).

**Conclusion:** The study demonstrates a substantial household burden of scabies and highlights the need for coordinated outbreak responses that prioritize household-level prevention (reducing sharing of personal items), improved access to effective treatment, and stronger health system–community engagement. Triangulation of regression results with community narratives supports AfyaData’s value for standardized, criteria-informed investigation and targeted public health action in Tanzania.

**Author Summary:** Scabies is a contagious skin disease caused by tiny mites that burrow into the skin and cause intense itching and rash. It spreads mainly through prolonged skin-to-skin contact and by sharing personal items such as clothes and bedding. Scabies is common in many low-resource settings, but local evidence is often limited, which makes it difficult to plan effective prevention and outbreak response.

We investigated a scabies outbreak that occurred in Nyasa District, southern Tanzania, in September 2022. We interviewed 198 households in four villages using a mobile data collection tool (AfyaData) and a standardized questionnaire informed by international scabies guidance. We found a high household burden of scabies experience. Quantitative analysis showed that households reporting scabies were more likely to report sharing personal items and seeking/receiving treatment during the outbreak, and they also reported weaker collaboration with healthcare personnel. Open-ended responses supported these patterns: participants described the importance of washing clothes and bedding, concerns that medicines were ineffective or difficult to access, and a need for health worker outreach, education, and follow-up. Some responses reflected uncertainty and misconceptions about the cause of illness.

Our findings show that scabies outbreak control in rural settings requires household-focused prevention, timely access to effective treatment, and stronger coordination between communities and health services. The study also demonstrates how digital tools can support standardized outbreak investigation and guide targeted public health action aligned to control scabies and other neglected tropical diseases.

## 1. Introduction

Scabies is a highly contagious skin infection caused by the mite Sarcoptes scabiei var. hominis, which burrows into the skin and lays eggs, resulting in intense itching that leads to skin lesions. Transmission occurs primarily through prolonged skin-to-skin contact. Scabies can affect people of any age, gender, race, or income level worldwide. (1) However, it is most common in low-income tropical areas, where children and older adults are susceptible (2). Untreated scabies can cause skin breakdown and secondary bacterial infections, which can lead to serious problems such as septicemia, rheumatic heart disease, and kidney disorders. (3). (4). Annually, scabies affects roughly 250 million people around the world (1). In Africa, scabies continues to be a significant public health issue among school-aged children, with a pooled prevalence estimated at 10.8%, suggesting an increasing trend in recent years, highlighting ongoing transmission and insufficient control measures in endemic areas (5) In 2022, Nyasa District experienced a major outbreak of suspected scabies in Chiwanda Ward, first reported in Mtupale village and subsequently affecting numerous hamlets and households. According to (6, 7), the incident developed with a significant portion of affected individuals remaining at home due to geographic barriers to health facilities. The outbreak offered a distinctive opportunity to conduct a retrospective analysis of factors associated with scabies transmission in a rural Tanzanian environment. Comprehending the behavioral, healthcare-related, and spatial features of the outbreak is crucial for guiding enhanced surveillance, targeted interventions, and preparedness strategies for scabies and other neglected tropical diseases.

## 2. Materials and Methods

### 2.1 Study Site

The study was conducted in Chiwanda Ward, Nyasa District Council, which comprises four villages: Mtupale, Kwambe, Ngombo, and Chimate. The ward is in the Lake Nyasa zone, where livelihoods are mainly smallholder farming and fishing, which informed the sampling frame for this study.

**Figure 1.**
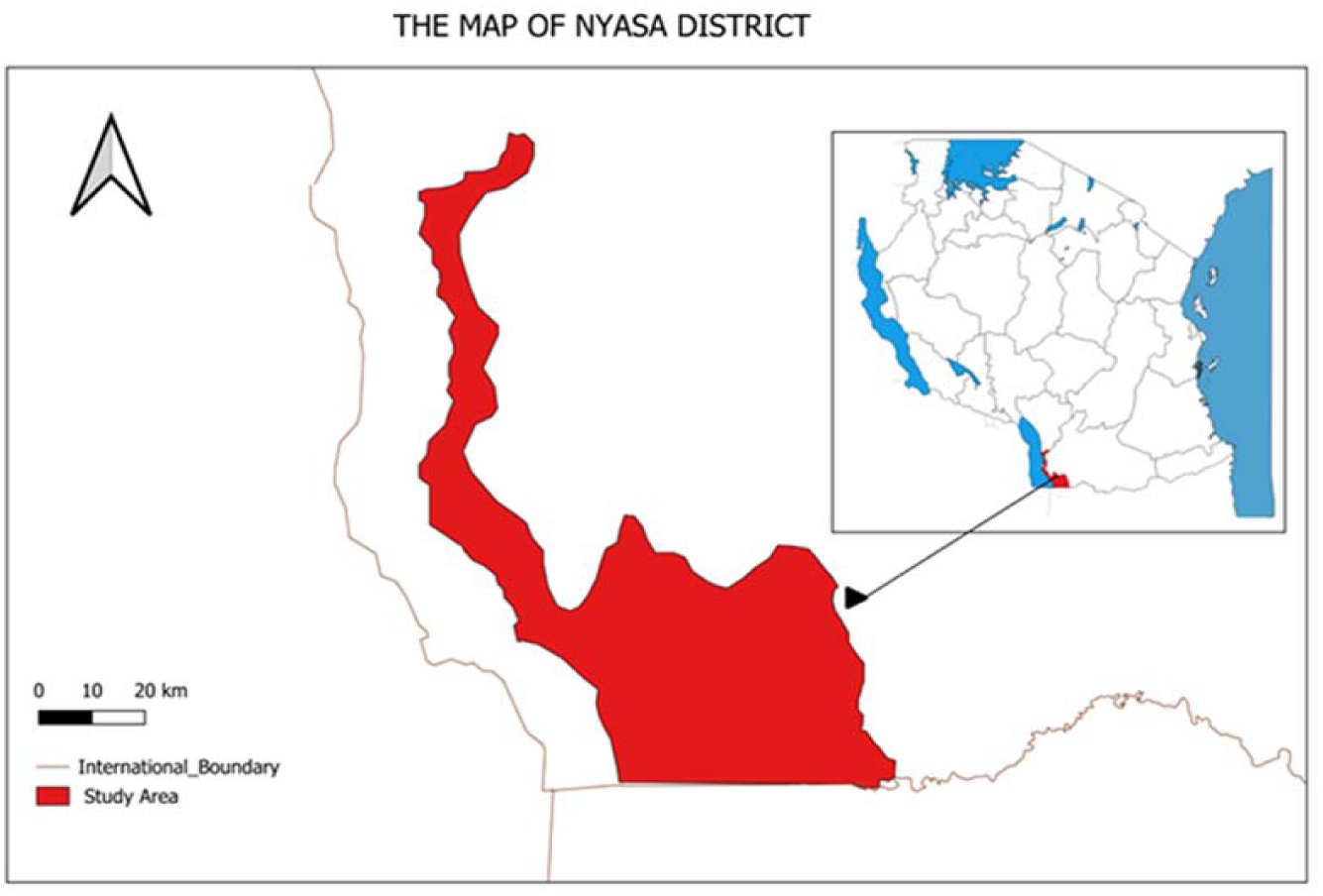
**Map of the study site**

### 2.2 Study Design and Population

The study employed a retrospective community-based cross-sectional analytical design, using the household as the unit of analysis. The study was conducted in Chiwanda Ward and included four village clusters. A total of 198 households were selected randomly, and one eligible adult respondent per household was interviewed to provide household-level information.

#### 2.2.1 Inclusion criteria

i. The household was occupied in the study villages during the September 2022 outbreak period.
ii. Written informed consent was obtained from all adult participants (≥18 years). For participants aged <18 years, written informed consent was obtained from a guardian where appropriate; assent was also obtained from the child. Participation was voluntary, and confidentiality was maintained. Photographs were taken only with specific consent and in compliance with NIMR ethical guidance.
iii. The respondent could reliably report household conditions, behaviors, and illness history during September 2022.

#### 2.2.2 Exclusion criteria

Households established after the outbreak period, households whose eligible respondent was not resident during the outbreak period, and households that declined participation.

#### 2.2.3 Outcome definition

The primary outcome was household scabies experience during the September 2022 outbreak. A household was classified as “Yes” if the respondent reported that at least one household member had scabies-related symptoms during the outbreak, defined by syndromic features typical of scabies. To improve consistency and reduce misclassification with other common dermatoses, the symptom-based outcome definition and questionnaire items were refined with dermatology expert input and aligned with standardized diagnostic principles consistent with the International Alliance for the Control of Scabies (IACS) 2020 criteria (16).

#### 2.2.4 Household mapping and spatial description

Global Positioning System (GPS) coordinates were recorded for each visited household in support of spatial visualization of the outbreak distribution. Two household distribution maps were generated. Household mapping enabled description of the geographic spread of visited households, assessment of possible spatial clustering and heterogeneity in risk, and identification of areas with potential transmission concentration. The mapping provided contextual evidence to support the interpretation of household-level environmental exposures and response practices in relation to the observed outbreak pattern.

#### 2.4.5 Sampling and Sample Size Estimation

Chiwanda Ward was purposively selected from the 20 wards of Nyasa District because it was the source of the first reported signal of a pruritic rash during the September incident, identified through the Event-Based Surveillance (EBS) system (Nyasa District Health Office, 2022a). The four villages in the ward, Mtupale, Kwambe, Ngombo, and Chimate, were treated as sampling clusters to ensure geographic coverage and to reflect the natural clustering of scabies transmission within communities and households. Households were selected by random sampling from each village. Data collectors, supported by Community Health Workers (CHWs) for community entry and household identification, visited selected households, explained the study, and obtained written informed consent before the interview. One eligible adult respondent was interviewed per household.

### 2.5 Sample size determination

The sample size for the retrospective community-based cross-sectional household survey was determined using a precision-based approach, which is appropriate when the key objective is to estimate household proportions within a pre-specified margin of error, rather than powering a single hypothesis test. (17)

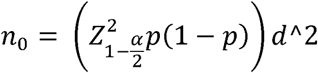

Where n_0 is_ the initial required sample size, Z_(1-α/2) is the standard normal value for a two-sided confidence level, p is the anticipated household proportion, and d is the desired precision. (18). Because no reliable ward-level estimate for the household proportion was available, a conservative value of p=0.50 was used to maximize the sample size and protect precision. With 95% confidence (Z=1.96) and ±7% precision (d=0.07). (19–21). Therefore, a minimum of 196 households was required. In the field, 198 households were successfully interviewed, which exceeds the minimum size needed and maintains the intended precision for estimating key household proportions.

### 2.6 Data Collection Procedure

Before field implementation, the study team configured the AfyaData mobile platform (22,23) to support the household survey. With technical support from SACIDS One Health, the structured questionnaire was digitized, uploaded to AfyaData, and tested for functionality. To improve content validity and minimize misclassification with other common dermatoses, the tool was reviewed and refined with input from a dermatology expert, ensuring alignment with syndromic features. The AfyaData application was installed and configured on smartphones used by data collectors, including user account setup, loading of the finalized form, and verification of GPS capture. Data collectors received practical orientation on interview procedures, consent administration, and use of the mobile tool, including troubleshooting and data synchronization steps. Field data collection was conducted at the household level in Chiwanda Ward across the four selected villages. For each sampled household, the field team visited the home, introduced the study with support from CHWs to facilitate community entry and household identification, obtained written informed consent, and interviewed one eligible adult per household, preferably the household head or primary caregiver, to provide household-level information. Interviews were conducted face-to-face using the digitized questionnaire, capturing socio-demographic characteristics, history of itching and skin rash during the outbreak period, household living and environmental conditions, hygiene practices, treatment history, and health-seeking behaviors, in line with the study objectives. To standardize recall and reduce variability across respondents, all questions were anchored specifically to the September 2022 outbreak period. Responses were entered directly into AfyaData at the point of interview, with built-in validation checks supporting completeness and internal consistency. GPS coordinates were recorded for each household to enable household mapping and support spatial epidemiological analyses.

### 2.7 Data analysis

Data from AfyaData were exported in Excel, checked for completeness and consistency, coded, and analyzed using SPSS version 23. The unit of analysis was the household, and the final analytic dataset comprised 198 households drawn from the four study villages. The outcome variable was a household with self-reported scabies symptoms during the September 2022 outbreak period, based on the standardized household interview responses. Descriptive statistics were used to summarize household socio-demographic, environmental, behavioral, and health-service related characteristics. At the bivariate level, associations between each independent variable and household scabies experience were assessed using Pearson’s chi-square test. Variables with p ≤ 0.20 at bivariate analysis were screened for multivariable modelling, alongside variables considered epidemiologically important a priori based on literature and field context. A binary multivariable logistic regression model was then fitted to estimate adjusted associations between predictors and household scabies experience. Model building used the Backward Likelihood Ratio (Backward LR) approach: an initial model containing all candidate variables was specified, and the least significant variables were removed sequentially using the likelihood ratio test until a parsimonious final model was obtained. Based on the screening and a priori criteria, candidate variables included village, education level, awareness of the outbreak, previous history of scabies, frequency of sharing items, main household water source, animal bathing practices, treatment received during the outbreak, preventive measures communicated/implemented, and collaboration of healthcare personnel/organizations. The final retained predictors are presented in the results section. Multicollinearity was assessed using the Variance Inflation Factor (VIF), with VIF < 2.5 considered acceptable. Overall model adequacy was assessed using the Omnibus Likelihood Ratio test, and goodness-of-fit was evaluated using the Hosmer–Lemeshow test. Results are reported as Adjusted Odds Ratios (AORs) with 95% Confidence Intervals (CI). Statistical significance was set at p < 0.05.

### 2.8 Ethical Consideration

The research clearance and ethical protocols of this study were approved by the Sokoine University of Agriculture and given reference numbers *SUA/ADM/R.1/8/1168* and DPRTC/SUA/000000, respectively. The permission to conduct this study in Ruvuma, Nyasa, was granted by the National Institute for Medical Research, reference number *NIMR/HQ/R.8a/Vol.IX/4583*.

## 3 Results

### 3.1 Socio-Demographic Characteristics of the Study Population (n = 198)

A total of 198 households were surveyed from four villages in Chiwanda Ward: Chimate (n=50; 25.3%), Kwambe (n=48; 24.2%), Mtupale (n=50; 25.3%), and Ngombo (n=50; 25.3%). Overall, 54.0% (n=107) of respondents were female. The median age of respondents was 46 years (IQR 34–56). Most respondents had primary education (71.7%; n=142), while 18.7% (n=37) reported no formal education and 9.6% (n=19) had secondary education or higher. Regarding occupation, 50.5% (n=100) were crop farmers, followed by fishermen/livestock keepers (25.3%; n=50) and business/formally employed (20.7%; n=41). Most respondents identified as Christians (96.0%; n=190). In terms of marital status, 57.6% (n=114) were in monogamous marriages. (Table 1)

**Table 1:** Socio-demographic characteristics of the study population.

| *Characteristic* | *Category* | *N* | *%* |
| --- | --- | --- | --- |
| Village name | Chimate | 50 | 25.3 |
|  | Kwambe | 48 | 24.2 |
|  | Mtupale | 50 | 25.3 |
|  | Ngombo | 50 | 25.3 |
| Sex | Female | 107 | 54.0 |
|  | Male | 91 | 46.0 |
| Age | 18-29 | 19 | 9.6 |
|  | 30-39 | 44 | 22.2 |
|  | 40-49 | 48 | 24.2 |
|  | 50-59 | 47 | 23.7 |
|  | 60 | 40 | 20.2 |
| Education Level | No education | 37 | 18.7 |
|  | Primary | 142 | 71.7 |
|  | High school and above | 19 | 9.6 |
| Occupation | Business/employed | 41 | 20.7 |
|  | crop farmer | 100 | 50.5 |
|  | fisherman/livestock keeper | 50 | 25.3 |
|  | None/retired | 7 | 3.5 |
| Religion | Christian | 190 | 96.0 |
|  | Muslim/other | 8 | 4.0 |
| Marital Status | Divorced/Separated | 24 | 12.1 |
|  | Married (Monogamy) | 114 | 57.6 |
|  | Married (polygamy) | 14 | 7.1 |
|  | Single | 19 | 9.6 |
|  | Widow/widower | 27 | 13.6 |

### 3.2 Scabies outbreak awareness and household scabies experience

Overall, 88.9% (n=176) reported being aware of a scabies outbreak in their community. Among those aware, first awareness was most commonly reported in August (17.0%) and December (14.8%), and awareness was relatively distributed across quarters (Q1–Q4). In total, 60.6% (n=120) reported that they or a household member experienced scabies during the outbreak period. Village-level household scabies prevalence showed clustering, with the highest prevalence in Mtupale (74.0%) and Chimate (72.0%), compared with Ngombo (50.0%) and Kwambe (45.8%). (Table 2A)

**Table 2A:** Outbreak awareness and household scabies experience (n=198 unless stated)

| *Characteristic* | *Category* | *N* | *%* |
| --- | --- | --- | --- |
| Aware of the outbreak in the village | No | 22 | 11.1 |
|  | Yes | 176 | 88.9 |
| Month of First Awareness | January | 7 | 4.0 |
|  | February | 21 | 11.9 |
|  | March | 20 | 11.4 |
|  | April | 18 | 10.2 |
|  | May | 10 | 5.7 |
|  | June | 9 | 5.1 |
|  | July | 11 | 6.3 |
|  | August | 30 | 17.0 |
|  | September | 4 | 2.3 |
|  | October | 8 | 4.5 |
|  | November | 12 | 6.8 |
|  | December | 26 | 14.8 |
| Period of awareness | Q1 (Jan-Mar) | 48 | 27.3 |
|  | Q2 (Apr-Jun) | 37 | 21.0 |
|  | Q3 (Jul-Sep) | 45 | 25.6 |
|  | Q4 (Oct-Dec) | 46 | 26.1 |
| Household Scabies Experience | No | 78 | 39.4 |
|  | Yes | 120 | 60.6 |
| Previous history of scabies | No | 115 | 58.1 |
|  | Not sure | 25 | 12.6 |
|  | Yes | 58 | 29.3 |

### 3.3 Clinical pattern and within-household transmission (affected households, n=120)

Among affected households, scabies was commonly reported on wrists (77.5%), fingers (78.3%), elbows (63.3%), and toes (60.0%), indicating a typical distribution consistent with close contact and household spread. The median number of body sites reported per affected household was 4 sites (IQR 3–6), and 77.5% reported ≥3 body sites, suggesting substantial symptom burden. (Tables 2B–2C). Within affected households, the median number of affected members was 2 (IQR 1–3), with a median household size of 6 (IQR 5–8). This corresponds to a median within-household attack proportion of 25% (IQR 20%–42.9%). In households where age-disaggregated counts were recorded (n=41 households), children under 5 years contributed 37.3% of affected individuals, suggesting meaningful involvement of young children in household transmission (Table 2D).

**Table 2B:** Body sites affected among scabies-affected households (n=120)

| Body site (among affected households, n=120) | N | % |
| --- | --- | --- |
| Wrists | 93 | 77.5 |
| Fingers | 94 | 78.3 |
| Elbows | 76 | 63.3 |
| Toes | 72 | 60.0 |
| Buttocks | 65 | 54.2 |
| Genitals | 64 | 53.3 |
| Armpits | 60 | 50.0 |

**Table 2C:**
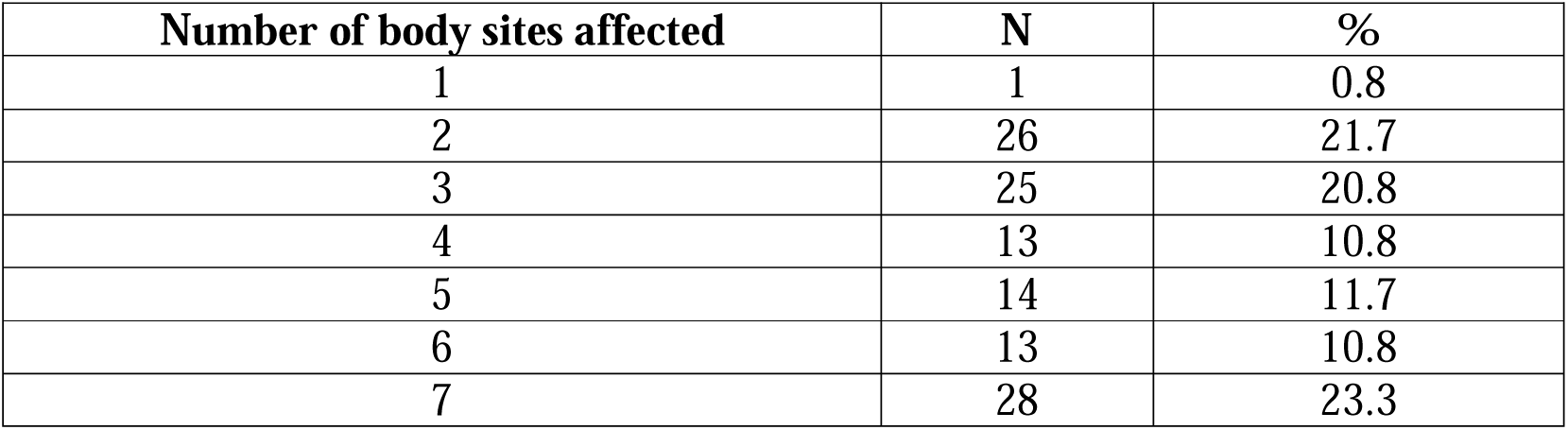
Number of body sites affected per household (affected households, n=120)

**Table 2D:** Number of affected members per household (affected households, n=120)

| Affected members in household (among affected households, n=120) | N | % |
| --- | --- | --- |
| 1 | 49 | 40.8 |
| 2 | 25 | 20.8 |
| 3 | 13 | 10.8 |
| 4 | 7 | 5.8 |
| 5 | 4 | 3.3 |
| 6 | 8 | 6.7 |
| 7 | 7 | 5.8 |
| 8 | 7 | 5.8 |

### 3.4 Household health and hygiene experience (n=198)

Item-sharing (towels/clothes/bedding) was reported at least rarely in 34.3%, including 16.2% who shared several times weekly/daily. Most households reported twice-daily bathing (69.7%), and normal (non-medicated) soap (85.4%) was most common. Water sources were mixed (multiple responses): RUWASA water (73.2%) was common, but 26.8% did not report RUWASA and relied on other sources, including springs, unprotected wells, and lake water. Domestic animals were kept by 72.2%, and among animal-keeping households, 65.0% reported never bathing animals. (Table 3) Note: The weakened immunity variable had 2 missing responses.

**Table 3:**
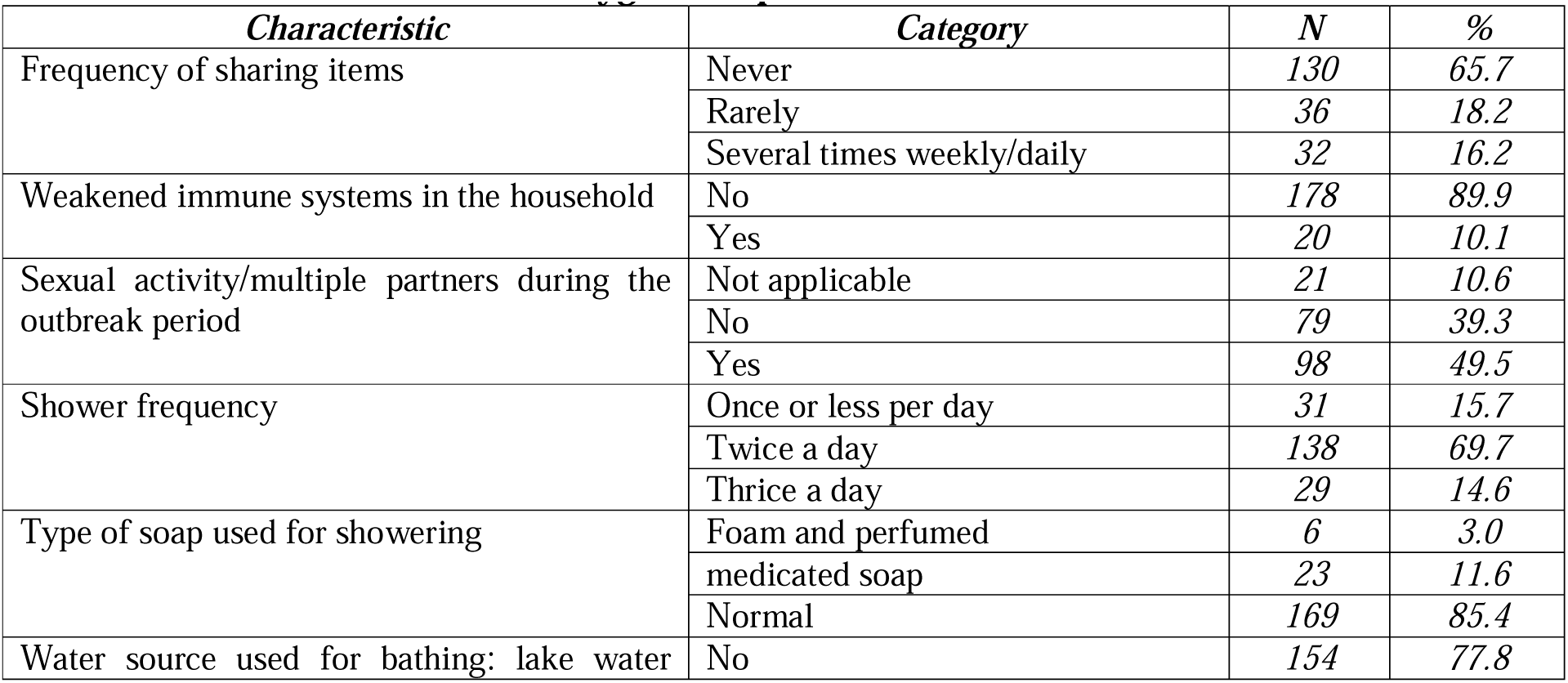
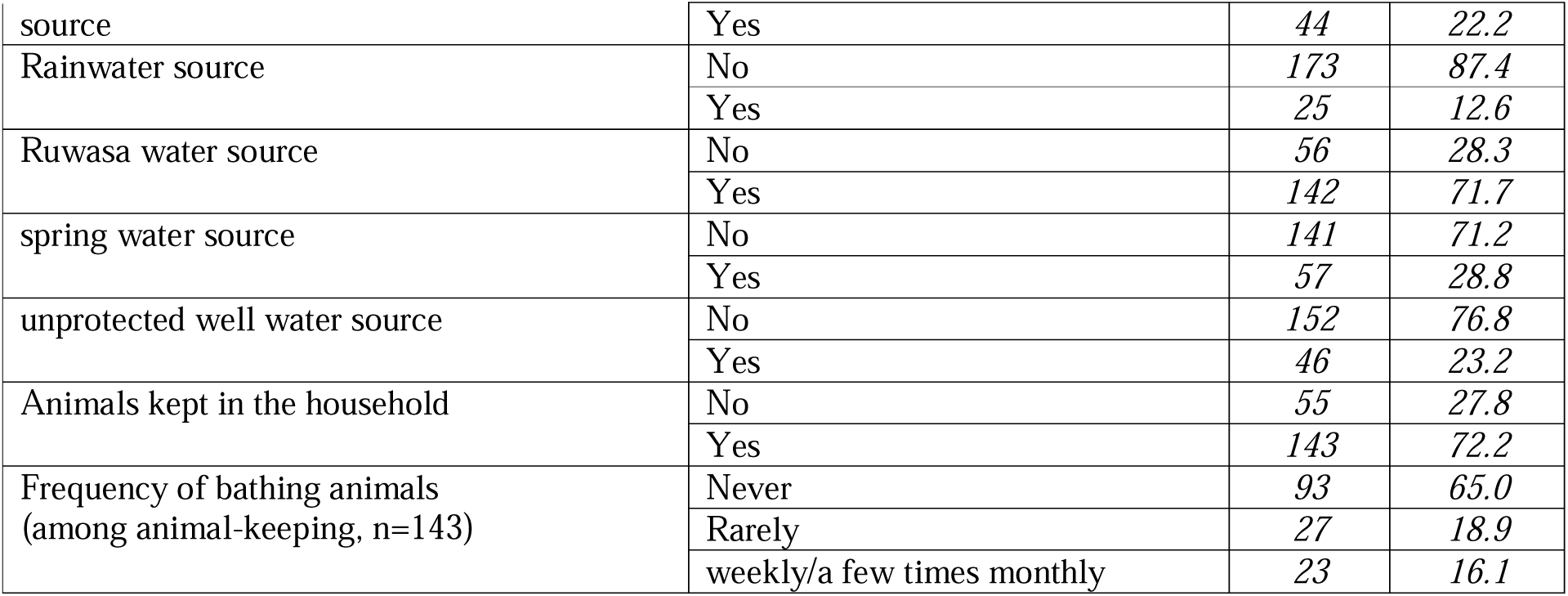
Household Health and Hygiene Experience.

### 3.5 Scabies treatment and control measures (n=198)

Overall, 52.0% (n=103) reported that specific treatment was provided, while 35.4% reported no treatment and 12.6% were unsure. Among treated affected households (n=78), 38.5% reported combined oral + topical regimens, 37.2% oral only, and 24.4% topical only, suggesting heterogeneous treatment practices. Preventive measures were reported by 45.5% (n=90); among these, prevention largely involved health education/personal hygiene (95.6%), followed by isolation of affected individuals (62.2%) and cleaning/disinfection (33.3%). Reported collaboration with healthcare personnel/organizations was 42.9%, while 43.4% reported no collaboration. (Tables 4A–4C)

**Table 4A:** Treatment and outbreak response measures (n=198)

| *Characteristic* | *Category* | *N* | *%* |
| --- | --- | --- | --- |
| Specific treatment provided during the outbreak | *No* | 70 | 35.4 |
|  | *not sure* | 25 | 12.6 |
|  | *Yes* | 103 | 52.0 |
| Preventive measures communicated/implemented | *No* | 81 | 40.9 |
|  | *not sure* | 27 | 13.6 |
|  | *Yes* | 90 | 45.5 |
| collaboration of healthcare personnel/organizations | *No* | 86 | 43.4 |
|  | *not sure* | 27 | 13.6 |
|  | *Yes* | 85 | 42.9 |

**Table 4B:** Reported treatment regimen among treated affected households (n=78)

| Medication regimen (among treated affected households, n=78) | N | % |
| --- | --- | --- |
| Oral + topical | 30 | 38.5 |
| Oral only | 29 | 37.2 |
| Topical only | 19 | 24.4 |

**Table 4C:** What prevention meant in practice.

| Component | N among prevention=Yes (n=90) | % among prevention=Yes | N among affected + prevention = yes (n=58) | % among affected + prevention= Yes |
| --- | --- | --- | --- | --- |
| Health education/personal hygiene | 86 | 95.6 | 55 | 94.8 |
| Isolation of affected individuals | 56 | 62.2 | 38 | 65.5 |
| Cleaning/disinfection of living space | 30 | 33.3 | 26 | 44.8 |
| Administration of oral medication | 20 | 22.2 | 15 | 25.9 |
| Use of topical treatment | 26 | 28.9 | 17 | 29.3 |

### 3.6 Factors associated with self-reported scabies (univariable and multivariable)

In univariable logistic regression, village clustering was evident: compared with Ngombo, households in Chimate (OR=2.57; p=0.026) and Mtupale (OR=2.85; p=0.015) had higher odds of reported scabies. Experience/awareness factors were strongly related to case reporting: being aware of the outbreak was associated with higher odds of reporting scabies (OR=6.54; p<0.001). Behavioral/environmental signals were also important: compared with never sharing items, rare item-sharing had markedly higher odds (OR=4.85; p=0.001), and frequent sharing also increased odds (OR=2.91; p=0.016). Not reporting RUWASA water use was associated with higher odds (OR=2.50; p=0.011), and among animal-keeping households, never bathing animals increased odds (OR=4.58; p=0.002). (Tables 5–7). Treatment variables showed reverse-direction patterns consistent with response targeting: households reporting treatment were more likely to be scabies-affected (OR=2.95; p=0.001), indicating treatment followed illness rather than causing it. (Table 8)

| Characteristic | Category | No scabies | No % | Scabies | Scabies % | OR | 95% CI | p-value |
| --- | --- | --- | --- | --- | --- | --- | --- | --- |
| Village name | Chimate | 14 | 28.0 | 36 | 72.0 | 2.571 | 1.122–5.895 | 0.026 |
|  | Kwambe | 26 | 54.2 | 22 | 45.8 | 0.846 | 0.383–1.871 | 0.68 |
|  | Mtupale | 13 | 26.0 | 37 | 74.0 | 2.846 | 1.228–6.597 | 0.015 |
|  | Ngombo | 25 | 50.0 | 25 | 50.0 | Ref |  |  |
| Gender | Female | 41 | 38.3 | 66 | 61.7 | 1.103 | 0.623–1.954 | 0.737 |
|  | Male | 37 | 40.7 | 54 | 59.3 | Ref |  |  |
| Education level | No education | 17 | 45.9 | 20 | 54.1 | 1.307 | 0.431–3.961 | 0.636 |
|  | Primary | 51 | 35.9 | 91 | 64.1 | 1.983 | 0.756–5.197 | 0.164 |
|  | Secondary and above | 10 | 52.6 | 9 | 47.4 | Ref |  |  |
| Occupation | Business/Employed | 16 | 39.0 | 25 | 61.0 | Ref |  |  |
|  | Crop farmer | 39 | 39.0 | 61 | 61.0 | 1.001 | 0.475–2.109 | 0.998 |
|  | Fisherman/Livestock keeper | 21 | 42.0 | 29 | 58.0 | 0.884 | 0.381–2.052 | 0.774 |
|  | None/Retired | 2 | 28.6 | 5 | 71.4 | 1.600 | 0.276–9.261 | 0.6 |
|  | Business/Employed | 16 | 39.0 | 25 | 61.0 | Ref |  |  |
| Religion | Christian | 76 | 40.0 | 114 | 60.0 | Ref |  |  |
|  | Muslim/other | 2 | 25.0 | 6 | 75.0 | 2.000 | 0.393–10.171 | 0.404 |
| Marital status | Divorced/Separated | 10 | 41.7 | 14 | 58.3 | 1.300 | 0.429–3.938 | 0.643 |
|  | Married (monogamy) | 42 | 36.8 | 72 | 63.2 | 1.592 | 0.684–3.707 | 0.281 |
|  | Married (polygamy) | 4 | 28.6 | 10 | 71.4 | 2.321 | 0.582–9.261 | 0.233 |
|  | Single | 9 | 47.4 | 10 | 52.6 | 1.032 | 0.319–3.341 | 0.958 |
|  | Widow/widower | 13 | 48.1 | 14 | 51.9 | Ref |  |  |

**Table 6:** Awareness/experience factors (univariable)

| Characteristic | Category | No scabies | No % | Scabies | Scabies % | OR | 95% CI | p-value |
| --- | --- | --- | --- | --- | --- | --- | --- | --- |
| Aware of the outbreak in the village | No | 17 | 77.3 | 5 | 22.7 | Ref |  |  |
|  | Yes | 61 | 34.7 | 115 | 65.3 | 6.541 | 2.255–18.971 | <0.001 |
| Period of first awareness (among aware) | Q1 | 19 | 39.6 | 29 | 60.4 | 1.174 | 0.516–2.669 | 0.702 |
|  | Q2 | 11 | 29.7 | 26 | 70.3 | 1.818 | 0.728–4.539 | 0.2 |
|  | Q3 | 11 | 24.4 | 34 | 75.6 | 2.378 | 0.971–5.822 | 0.058 |
|  | Q4 | 20 | 43.5 | 26 | 56.5 | Ref |  |  |
| Previous history of scabies | No | 44 | 38.3 | 71 | 61.7 | Ref |  |  |
|  | Not sure | 16 | 64.0 | 9 | 36.0 | 0.349 | 0.142–0.857 | 0.022 |
| Previous history of scabies | Yes | 18 | 31.0 | 40 | 69.0 | 1.377 | 0.704–2.695 | 0.35 |
| Previous history of scabies |  |  |  |  |  |  |  |  |

**Table 7:**
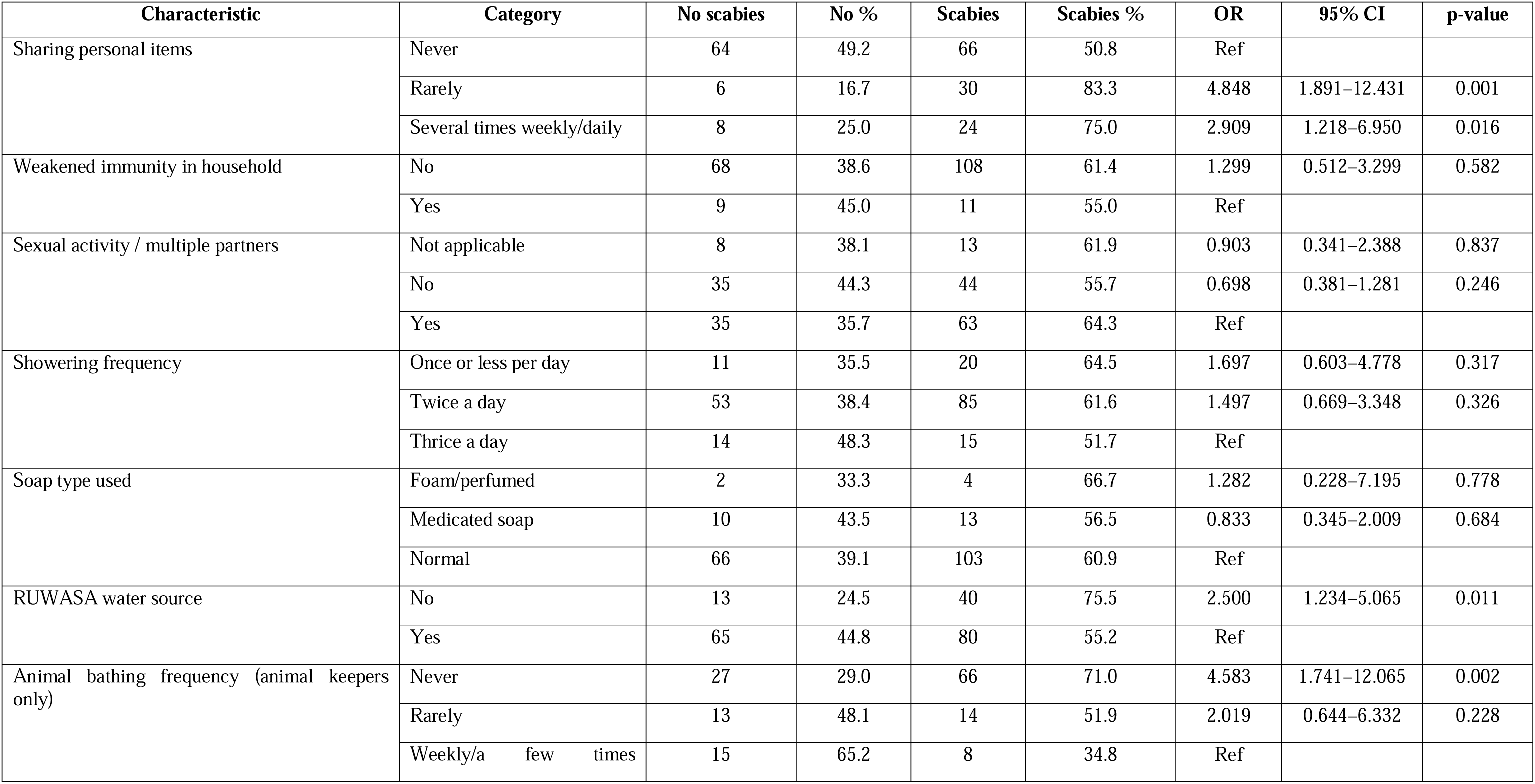
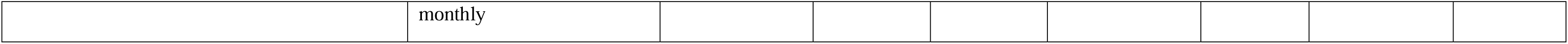
Hygiene/behavior factors (univariable)

**Table 8:** Treatment/control measures (univariable)

| Characteristic | Category | No scabies | No % | Scabies | Scabies % | OR | 95% CI | p-value |
| --- | --- | --- | --- | --- | --- | --- | --- | --- |
| Specific treatment provided | No | 34 | 48.6 | 36 | 51.4 | Ref |  |  |
|  | Not sure | 19 | 76.0 | 6 | 24.0 | 0.298 | 0.106–0.836 | 0.021 |
|  | Yes | 25 | 24.3 | 78 | 75.7 | 2.947 | 1.538–5.645 | 0.001 |
| Preventive measures implemented | No | 30 | 37.0 | 51 | 63.0 | Ref |  |  |
|  | Not sure | 16 | 59.3 | 11 | 40.7 | 0.404 | 0.166–0.985 | 0.046 |
|  | Yes | 32 | 35.6 | 58 | 64.4 | 1.066 | 0.571–1.990 | 0.841 |
| Collaboration with healthcare personnel/organizations | No | 31 | 36.0 | 55 | 64.0 | Ref |  |  |
|  | Not sure | 16 | 59.3 | 11 | 40.7 | 0.388 | 0.160–0.939 | 0.036 |
|  | Yes | 31 | 36.5 | 54 | 63.5 | 0.982 | 0.526–1.832 | 0.954 |

### 3.9 Multivariable model (parsimonious)

Variables with p ≤ 0.20 in univariable analysis and those considered epidemiologically important were entered into a binary multivariable logistic regression model to identify factors independently associated with household scabies experience during the September 2022 outbreak period. Adjusted odds ratios (AOR) with 95% confidence intervals (CI) were reported. To reduce the risk of overfitting, given the smaller non-case group and the number of candidate predictors, a parsimonious final model was retained. In the final model, households in which respondents were “not sure” about previous scabies history had lower odds of reporting scabies experience compared with households reporting no prior history (AOR = 0.119; 95% CI: 0.031–0.462; p = 0.002). This pattern likely reflects misclassification in retrospective reporting rather than a protective biological effect. Households reporting rare sharing of personal items had higher odds of scabies experience compared with those reporting never sharing (AOR = 4.233; 95% CI: 1.036–17.288; p = 0.044). Sharing items several times weekly/daily also showed elevated odds with borderline statistical evidence (AOR = 3.537; 95% CI: 0.986–12.685; p = 0.053). Households reporting that specific treatment was provided during the outbreak had higher odds of scabies experience compared with those reporting no treatment (AOR = 4.415; 95% CI: 1.310–14.881; p = 0.017), which is consistent with response targeting rather than treatment causing scabies. Finally, reporting no collaboration with healthcare personnel/organizations was associated with higher odds of scabies experience compared with reporting collaboration (AOR = 4.871; 95% CI: 1.391–17.060; p = 0.013), supporting the importance of coordinated outbreak response. Variables including village of residence, education level, period of awareness, RUWASA water source, and frequency of bathing animals did not retain independent associations after adjustment and were excluded from the final model, suggesting that their crude relationships were likely explained by more proximal behavioral and response-related factors. (Table 9)

**Table 9:** Multivariable logistic regression (final model)

| Characteristic | Category | AOR | 95% CI | p-value |
| --- | --- | --- | --- | --- |
| Previous history of scabies | No | Ref |  |  |
|  | Not sure | 0.235 | 0.078–0.710 | 0.01 |
|  | Yes | 1.271 | 0.575–2.805 | 0.553 |
| Sharing personal items | Never | Ref |  |  |
|  | Rarely | 4.059 | 1.467–11.226 | 0.007 |
|  | Several times<br>weekly/daily | 4.688 | 1.641–13.393 | 0.004 |
| Specific treatment provided | No | Ref |  |  |
|  | Not sure | 0.249 | 0.067–0.928 | 0.038 |
|  | Yes | 4.705 | 2.051–10.795 | <0.001 |
| Collaboration with healthcare<br>personnel/organizations | Yes | Ref |  |  |
|  | Not sure | 1.683 | 0.482–5.881 | 0.415 |
|  | No | 2.035 | 0.877–4.720 | 0.098 |

*Model performance: LR $\chi^2 \approx 56.3$ ( $p < 0.001$ ), McFadden pseudo- $R^2 \approx 0.21$ , AUC $\approx 0.80$ .*

**Table 10.** Thematic distribution of open-ended responses (multiple themes per response)

| Theme | Q24 (n=110) | Q25 (n=47) | Q26 (n=87) |
| --- | --- | --- | --- |
| Hygiene & personal cleanliness | 64 (58.2%) | 6 (12.8%) | 2 (2.3%) |
| Treatment effectiveness & adherence/medication concerns | 12 (10.9%) | 18 (38.3%) | 38 (43.7%) |
| Access/cost & availability of services/medicines | 8 (7.3%) | 7 (14.9%) | 31 (35.6%) |
| Health education & awareness | 8 (7.3%) | 3 (6.4%) | 15 (17.2%) |
| Coordination/government response & outreach | 3 (2.7%) | 19 (40.4%) | 16 (18.4%) |
| Isolation/avoid contact | 8 (7.3%) | 0 (0.0%) | 0 (0.0%) |
| Laundry, bedding & avoiding sharing items | 9 (8.2%) | 1 (2.1%) | 0 (0.0%) |
| Recurrence/treatment failure & need for sustained control | 3 (2.7%) | 0 (0.0%) | 0 (0.0%) |
| Traditional beliefs/home remedies/misconceptions | 3 (2.7%) | 1 (2.1%) | 0 (0.0%) |
| Early care-seeking & reporting | 1 (0.9%) | 2 (4.3%) | 0 (0.0%) |

*Note: Percentages use the non-empty responses for each question (Q24=110, Q25=47, Q26=87).*

**Table 11.**
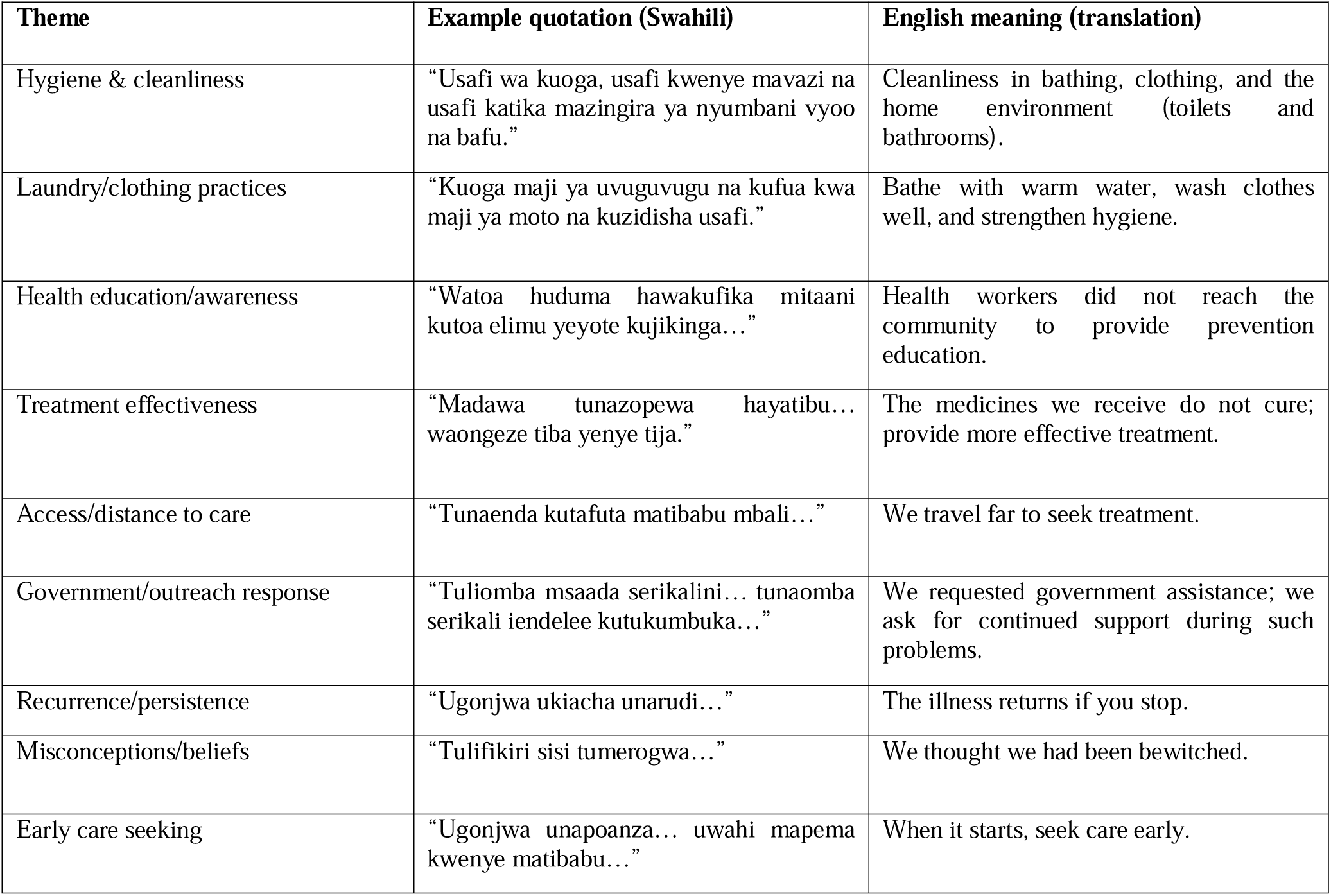
Illustrative quotations supporting key themes (translated)

**Table 12:** Matrix linking Table 9 factors to open-ended themes.

| Table 9 factor | Adjusted result | Matching open-ended theme(s) (Q24–Q26) | Example evidence (illustrative) |
| --- | --- | --- | --- |
| Sharing personal items | Rarely AOR=4.059;<br>Frequent AOR=4.688 | Laundry/bedding; avoid sharing items; clothing hygiene | “...kufua... na kuzidisha usafi” |
| Treatment provided | Yes, AOR=4.705. | Treatment effectiveness concerns; access/availability barriers | “madawa... hayatibu”;<br>“matibabu mbali” |
| Collaboration | No AOR=2.035<br>(p=0.098) | Outreach/coordination requests; lack of education/follow-up | “watoa huduma hawakufika...” |
| “Not sure” history/treatment | History AOR=0.235;<br>Treatment AOR=0.249 | Uncertainty/misconceptions | “tulifikiri... tumerogwa” |

Table 12: Matrix linking Table 9 factors to open-ended themes
| Table 9 factor | Adjusted result | Matching open-ended theme(s) (Q24–Q26) | Example evidence (illustrative) |
| --- | --- | --- | --- |
| Sharing personal items | Rarely AOR=4.059;<br>Frequent AOR=4.688 | Laundry/bedding; avoid sharing items; clothing hygiene | “...kufua... na kuzidisha usafi” |
| Treatment provided | Yes AOR=4.705 | Treatment effectiveness concerns; access/availability barriers | “madawa... hayatibu”; “matibabu mbali” |
| Collaboration | No AOR=2.035 (p=0.098) | Outreach/coordination requests; lack of education/follow-up | “watoa huduma hawakufika...” |
| “Not sure” history/treatment | History AOR=0.235;<br>Treatment AOR=0.249 | Uncertainty/misconceptions | “tulifikiri... tumerogwa” |

### 3.9 Community perspectives from open-ended questions (thematic results)

Open-ended responses were analyzed using thematic content analysis for three questions (Q24–Q26). Responses were first cleaned to remove blanks, then coded into recurring themes. Because responses frequently contained multiple ideas (e.g., hygiene + treatment + access), multiple themes were allowed per response, so percentages can sum to more than 100%. After excluding blank entries, 110/198 (55.6%) households provided a non-empty response to Q24 (lessons learned), 47/198 (23.7%) to Q25 (additional comments), and 87/198 (43.9%) to Q26 (treatment/medications/access). A proportion of responses were brief/unclear and were retained but categorized as “other/uncoded” during coding (Q24: 20.0%; Q25: 27.7%; Q26: 26.4%).

## 4 Discussion

This study describes a large household burden of self-reported scabies during the 2022 outbreak in Chiwanda Ward, with 60.6% (120/198) of households reporting scabies in at least one member. The outbreak showed geographic clustering (higher odds in Chimate and Mtupale in univariable analysis), a clinical distribution consistent with scabies (wrists/fingers/elbows/toes commonly affected), and strong evidence that household contact behaviours and response-system gaps shaped transmission. In the final multivariable model, the most important independent correlates of household scabies experience were sharing personal items, reporting that treatment was provided, lack of collaboration with healthcare personnel/organizations, and “not sure” responses on prior scabies history (likely indicating information bias rather than true protection).

### 4.1 Outbreak burden and village-level clustering

The high proportion of affected households indicates intense transmission at the community level. The higher odds observed in Chimate and Mtupale compared with Ngombo suggest localized transmission hotspots. This is epidemiologically plausible because scabies spreads most efficiently through prolonged close contact within households and close networks, and transmission tends to cluster where household mixing is dense and response coverage is uneven (WHO, 2023; CDC, 2024). Programmatically, these village differences argue for micro-targeted outbreak control, early case finding, and contact management in hotspot villages before spread becomes generalized.

### 4.2 Clinical pattern supports plausibility of the self-reported case definition

Among affected households, the most commonly reported sites, wrists (77.5%), fingers (75.8%), elbows (63.3%), and toes (60.0%), fit the expected distribution of common scabies lesions and support the plausibility of household reports. Standardized approaches such as the 2020 IACS criteria exist to improve comparability and reduce misclassification in scabies epidemiology, especially where laboratory confirmation is not feasible (Engelman et al., 2020).

Because your outcome was self/household-reported symptoms during a past outbreak period, some misclassification remains likely; however, the consistent body-site pattern strengthens confidence that many reports represented true scabies.

### 4.3 Within-household transmission intensity

Your additional epidemiologic descriptors from the 120 affected households show that scabies was not merely sporadic: the median number of affected members per affected household was 2, with a median within-household attack proportion around one-quarter. This supports the core biology and transmission route—scabies spreads efficiently among people with repeated close contact in the same residence, and household members are a key transmission unit (WHO, 2023; CDC, 2024). A key limitation, however, is that household size/affected counts were captured mainly among affected households, restricting direct comparison of crowding between affected and unaffected households. This should be acknowledged as a measurement limitation.

### 4.4 Sharing personal items as a behavioural driver

The most actionable behavioural determinant in your multivariable model was sharing personal items. Households reporting rare sharing had significantly higher odds of scabies compared with those reporting never sharing (AOR ≈ 4.23), and frequent sharing also showed elevated odds (borderline evidence). This finding is consistent with the notion that, during outbreaks, shared clothes, towels, and bedding can sustain transmission—either directly (through prolonged close contact that accompanies sharing) or indirectly via contaminated items, particularly when infestation intensity is high or when crusted scabies occurs (CDC, 2024; WHO, 2023).

Important nuance for your discussion: WHO notes that fomite transmission is less likely in common scabies but may be more important for crusted scabies, whereas public health guidance commonly still includes avoiding shared bedding/clothing as a practical outbreak control measure (WHO EMRO guidance). Interpretation can be framed as item sharing being a proxy for close household mixing and shared sleeping/linen practices, which are central to household propagation.

### 4.5 Awareness: strong association likely reflects reverse causation

Although awareness of the outbreak was high (88.9%), awareness was also strongly associated with scabies status in univariable analysis (OR ≈ 6.41). In outbreak epidemiology, this pattern often reflects reverse causation: households that experienced illness are more likely to notice, discuss, and recall the outbreak than unaffected households. This should be presented as an expected bias in retrospective self-reporting rather than a causal risk factor. Your “month/quarter of first awareness” not being associated with scabies status also supports the idea that awareness timing is not the primary driver of risk, but rather an experience-related marker.

### 4.6 WASH and animal-related variables

In univariable analysis, not reporting RUWASA water and never bathing animals among animal-keeping households were associated with higher odds of scabies. In the multivariable model, these did not remain independent correlates after accounting for more proximal behavioral response factors. Epidemiologically, this suggests confounding by village and household practices: water access and animal care may reflect broader living conditions, but scabies transmission is still primarily driven by prolonged close contact and incomplete contact treatment (WHO, 2023; CDC, 2024).

For discussion, you can still mention WASH and household environment as contextual contributors (e.g., affecting laundering frequency or living density), while being clear that your adjusted model supports behavioural contact pathways and response quality as the key drivers.

### 4.7 Treatment and response measures: interpreting “treatment provided” and collaboration

Reporting that treatment was provided during the outbreak was strongly associated with household scabies experience in both univariable and multivariable analysis (AOR ≈ 4.42). This should be interpreted as response targeting: treatment was more likely delivered (or remembered) in households already affected, rather than treatment increasing risk. Cross-sectional retrospective designs commonly produce this pattern because exposure and outcome are assessed together, and interventions are often delivered in reaction to illness.

Your results also show important gaps in outbreak control:

- Only about half reported receiving specific treatment (52.0%).
- Preventive measures were reported by 45.5%, while 40.9% reported none.
- Reported collaboration with healthcare personnel/organizations was 42.9%, and 43.4% reported no collaboration.

These gaps matter because scabies control generally requires coordinated case detection plus treatment of cases and close contacts, and fragmented responses increase reinfestation and persistence (CDC, 2024; WHO EMRO RCCE guidance).

Your multivariable finding that lack of collaboration was associated with higher odds (AOR ≈ 4.87) supports the interpretation that outbreak control was not only biomedical (drugs) but also system/coordination-dependent.

### 4.8 Linking multivariable results (Table 9) with open-ended themes (Q24–Q26)

To support interpretation of the multivariable model, themes from open-ended responses (Section 3.10) were compared with the independent factors retained in Table 9. Overall, open-ended responses contained recurring content that corresponded to the main adjusted associations. The Table 9 association with sharing personal items (rare sharing AOR=4.059; frequent sharing AOR=4.688) corresponded with Q24 themes emphasizing laundry/bedding practices and avoiding sharing items, and broader messages around clothing and household cleanliness. Respondents described strengthening hygiene and laundering practices as key lessons, including statements such as “…kufua… na kuzidisha usafi.”. The Table 9 association with reported treatment provided (AOR=4.705) corresponded with Q26 and Q25 themes on treatment effectiveness/medication concerns and access/availability barriers. Respondents frequently reported that medicines were not effective or were insufficient and highlighted distance or service barriers (e.g., “madawa… hayatibu”; “tunaenda kutafuta matibabu mbali”).

The elevated adjusted odds for no collaboration (AOR=2.035; p=0.098) corresponded with Q25 and Q26 themes calling for stronger outreach, follow-up, and coordination from health personnel/government, including comments that health staff did not reach communities with education or follow-up. The inverse associations observed for “not sure” categories (previous history not sure AOR=0.235; treatment not sure AOR=0.249) corresponded with a smaller set of open-ended responses indicating uncertainty or misconceptions about the condition (e.g., attributing illness to non-medical causes), supporting the presence of information/recognition challenges in retrospective reporting.

### 4.8 Implications for scabies outbreak control in Nyasa District

Your findings point to a practical package aligned with current global scabies control thinking:

i. Household-centered response: prioritize treating cases and close contacts together, paired with clear messaging on avoiding item sharing during outbreaks (CDC, 2024; WHO EMRO guidance).
ii. Hotspot targeting: prioritize early action in high-prevalence villages (e.g., Mtupale/Chimate) to interrupt transmission networks.
iii. Strengthen collaboration: formalize coordination between facilities, CHWs, and village leadership to improve coverage, consistency, and follow-up.
iv. Consider community-wide approaches when indicated: evidence from island/community trials shows that ivermectin-based mass drug administration (MDA) can markedly reduce scabies prevalence and impetigo burden, supporting its use in appropriate endemic/outbreak contexts with strong implementation capacity (Romani et al., 2015; Romani et al., 2019).
v. Align with NTD roadmap direction: scabies is now included within the WHO NTD agenda and remains underreported in many settings, reinforcing the need to integrate scabies into routine plans and surveillance packages (WHO NTD Roadmap 2021–2030; WHO Executive Board report 2026).

### 4.9 Strengths and limitations

Strengths

i. Community-based design captured household-level behaviours and response experiences that facility data often miss.
ii. Symptom distribution aligns with known scabies patterns, supporting the plausibility of case classification.

**Limitations (and where you should explicitly justify)**

i. Self-reported retrospective outcomes can cause misclassification and recall bias; consider referencing standardized diagnostic frameworks such as IACS criteria as a benchmark (Engelman et al., 2020).
ii. Reverse causation likely explains associations such as “awareness” and “treatment provided” appearing as risk markers.
iii. Incomplete denominators for some household composition variables (recorded mainly among affected households) limit inference on crowding/household size as risk factors.
iv. Social desirability bias may affect reported hygiene behaviours (e.g., bathing frequency).
v. Multiple-response WASH variables can reflect village-level infrastructure differences, complicating causal interpretation.

## Conclusion

The 2022 scabies outbreak in Chiwanda Ward was characterized by high household burden, village clustering, and transmission patterns consistent with scabies spread through close contact and shared household practices. After adjustment, sharing personal items remained the strongest behavioral correlate, while treatment provided likely reflected response targeting rather than causation. Collaboration showed a suggestive protective direction but was not statistically conclusive after adjustment. Strengthening household-centered prevention messaging, standardized treatment of cases and contacts, and coordinated outbreak responsewith consideration of MDA in high-prevalence settings follow directly from your results and align with global scabies/NTD guidance.

## Supporting information

Supplemental 1 and will be used for the link to the file on the preprint site.

Supplemental 2, and will be used for the link to the file on the preprint site.

Supplemental Table 3, and will be used for the link to the file on the preprint site.

Supplemental 5, and will be used for the link to the file on the preprint site.

## 5. Acknowledgments

The authors gratefully acknowledge the Southern African Centre for Infectious Disease Surveillance (SACIDS Foundation for One Health) for providing invaluable technical support and access to the AfyaData digital surveillance platform, which was instrumental to this study. We extend our sincere gratitude to the Southern Highlands Community Health (SOHICOHE) organization for the support that made the field activities possible. We are deeply grateful to the community health workers and local leaders in Chiwanda ward whose tireless efforts in mobilizing participants and facilitating data collection were essential to the success of this research. Special thanks are due to the residents of Chimate, Kwambe, Mtupale, and Ngombo villages for their willing participation and cooperation. Ethical approval for this study was obtained from the National Institute for Medical Research, *NIMR/HQ/R.8a/Vol.IX/4583*, and permission to conduct the study was granted by the Nyasa District Council. Written informed consent was obtained from all study participants before data collection, ensuring full adherence to ethical research standards.

## 6. Data Availability Statement

The data supporting the findings of this study are derived from the Nyasa District Event-Based outbreak investigation report and household-level data collected using the AfyaData digital platform. The AfyaData dataset is securely stored on the AfyaData server under the management of the SACIDS Foundation for One Health. Due to ethical and confidentiality considerations, the raw household-level data are not publicly available but may be accessed upon reasonable request to the corresponding author and with approval from the Nyasa District Council and the National Institute for Medical Research (NIMR). Aggregated results supporting the conclusions of this study are included within the manuscript.

## Appendix 1 Supporting Document -S1

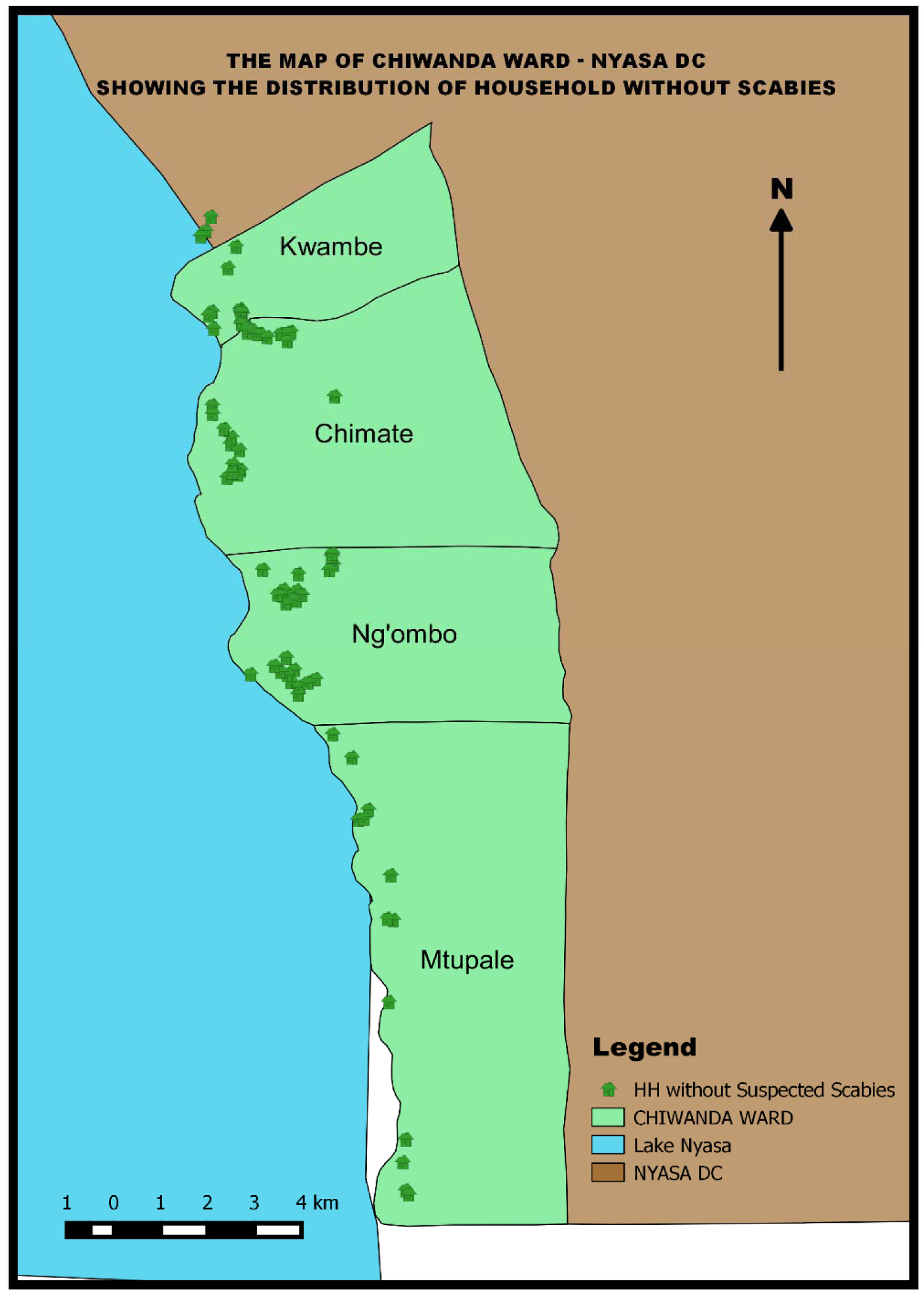
Spatial distribution of households without suspected scabies during the 2022 outbreak in Chiwanda Ward, Nyasa District. Green points represent households reporting no scabies symptoms in any household member (n=78; 39.4%). The map illustrates the geographic distribution of unaffected households across the four study villages, with relatively even dispersion across Kwambe and Ngombo villages. The blue area represents Lake Nyasa. The scale bar indicates 0-4 kilometers.

## Appendix 2 Supporting Document -S2

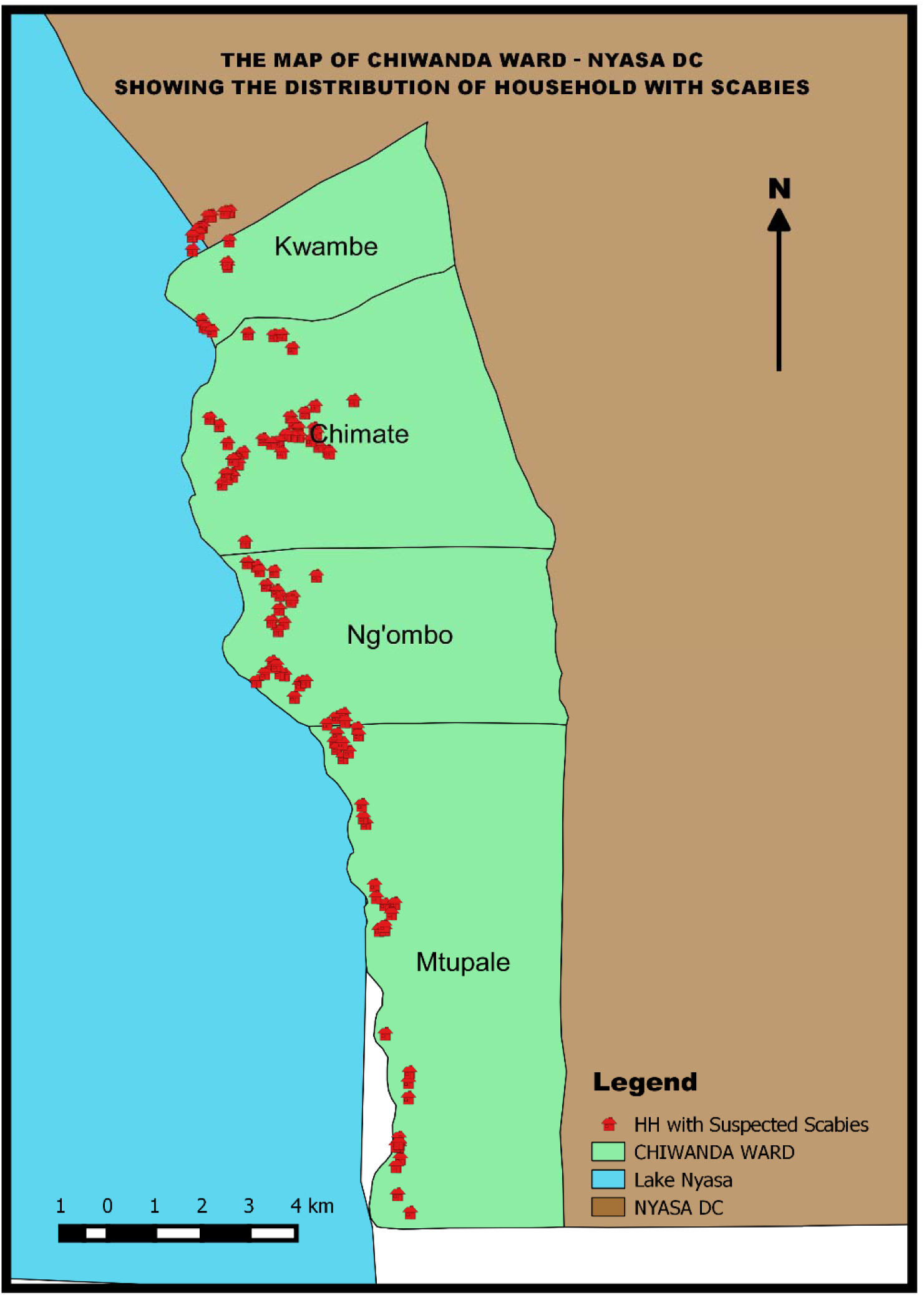
Red points represent households reporting scabies symptoms in the respondent or any household member (n=120; 60.6%). The map shows clustering patterns across four villages: Chimate, Kwambe, Mtupale, and Ngombo, with notable concentration in Chimate and Mtupale villages along the lakeshore. The blue area represents Lake Nyasa. The scale bar indicates 0-4 kilometers.

## References

1. Schneider S, Wu J, Tizek L, Ziehfreund S, Zink A. Prevalence of scabies worldwide—An updated systematic literature review in 2022. J Eur Acad Dermatol Venereol. 2023 Sept;37(9):1749–57.

2. El-Moamly AA. Scabies as a part of the World Health Organization roadmap for neglected tropical diseases 2021–2030: what we know and what we need to do for global control. Trop Med Health. 2021 Dec;49(1):64.

3. Matthewman J, Manago RZ, Dimessa Mbadinga LB, Šinkovec H, Völker K, Akinosho M, et al. A randomized controlled trial comparing the effectiveness of individual versus household treatment for Scabies in Lambaréné, Gabon. PLoS Negl Trop Dis. 2020;14(6): e0008423.

4. World Health Organization [Internet]. 2023 [cited 2026 Jan 9]. Scabies. Available from: https://www.who.int/news-room/fact-sheets/detail/scabies

5. Girma A, Abdu I, Teshome K. Prevalence and determinants of scabies among schoolchildren in Africa: A systematic review and meta-analysis. SAGE Open Med. 2024;

6. Nyasa District Health Office. Taarifa ya ugonjwa wa upele katika Kijiji cha Mtupale: Event-based surveillance verification report. Ruvuna, Nyasa District: Nyasa District Council; 2022 09 p. 5. (Weekly EBS Report). Report No.: Week 34.

7. Nyasa District Health Office. ALERT REGISTER NYASA DC - WEEK 34. Tanzania; 2022.

8. World Health Organization [Internet]. World Health Organization; 2021 Jan [cited 2026 Jan 16] p. 196. Available from: https://www.who.int/publications/i/item/9789240010352

9. Ministry of Health. Strategic master plan for the neglected tropical diseases control program: Tanzania Mainland (July 2021–June 2026) [Internet]. Ministry of Health (MoH), Tanzania; 2021 July. Available from: https://www.moh.go.tz/storage/app/uploads/public/

10. CDC. National Center for Emerging and Zoonotic Infectious Diseases (NCEZID). 2024 [cited 2026 Jan 11]. National Center for Emerging and Zoonotic Infectious Diseases (NCEZID). Available from: https://www.cdc.gov/ncezid/index.html

11. CDC. Scabies. 2025 [cited 2026 Jan 11]. Public Health Strategies for Scabies Outbreaks in Institutional Settings. Available from: https://www.cdc.gov/scabies/php/public-health-strategy/index.html

12. Ziebold C, Crane JS. Scabies. In StatPearls Publishing; 2025 [cited 2026 Jan 20]. Available from: https://www.ncbi.nlm.nih.gov/books/NBK544306/

13. Amoako YA, Phillips R, Arthur J, Abugri MA, Akowuah EK, Amoako KO, et al. A scabies outbreak in the North East Region of Ghana: The necessity for prompt intervention. PLoS Negl Trop Dis. 2020;

14. Balcha F, Bizuneh H, Hunduma F. Scabies Outbreak Investigation and Its Risk Factors in Gumbichu District, East Shewa Zone, Central Ethiopia: Unmatched Case-Control Study. 2022;

15. Kaburi BB, Ameme DK, Adu-Asumah G, Dadzie D, Tender EK, Addeh SV, et al. Outbreak of scabies among preschool children, Accra, Ghana, 2017. BMC Public Health. 2019 Dec;19(1):746.

16. Engelman D, Yoshizumi J, Hay RJ, Osti M, Micali G, Norton S, et al. The 2020 international alliance for the control of scabies consensus criteria for the diagnosis of scabies. Br J Dermatol. 2020;183(5):808–20.

17. Agresti A. The foundations of statistical science: A history of textbook presentations. Braz J Probab Stat. 2021 Nov [cited 2026 Jan 23];35(4). Available from: https://projecteuclid.org/journals/brazilian-journal-of-probability-and-statistics/volume-35/issue-4/The-foundations-of-statistical-science--A-history-of-textbook/10.1214/21-BJPS518.full

18. Johnston KM, Lakzadeh P, Donato BMK, Szabo SM. Methods of sample size calculation in descriptive retrospective burden of illness studies. BMC Med Res Methodol. 2019 Jan 9;19(1):9.

19. Wasserstein RL, Schirm AL, Lazar NA. Moving to a World Beyond “p < 0.05”. Am Stat. 2019 Mar 29;73(sup1):1–19.

20. Ahlbom A. Modern Epidemiology, 4th edition. TL Lash, TJ VanderWeele, S Haneuse, KJ Rothman. Wolters Kluwer, 2021. Eur J Epidemiol. 2021 July 3;36(8):767–8.

21. Rothman KJ, Huybrechts KF, Murray EJ. Epidemiology: an introduction. Oxford university press; 2024.

22. Karimuribo ED, Mutagahywa E, Sindato C, Mboera L, Mwabukusi M, Njenga MK, et al. A smartphone app (AfyaData) for innovative one health disease surveillance from community to national levels in Africa: intervention in disease surveillance. JMIR Public Health Surveill. 2017;3(4): e7373.

23. Mwabukusi M, Karimuribo ED, Rweyemamu MM, Beda E. Mobile technologies for disease surveillance in humans and animals. Onderstepoort J Vet Res. 2014 Apr 23;81(2):5 pages.

24. Engelman D, Marks M, Steer AC, Beshah A, Biswas G, Chosidow O, et al. A framework for scabies control. PLoS Negl Trop Dis. 2021;15(9): e0009661.

25. El-Moamly AA. Scabies as a part of the World Health Organization roadmap for neglected tropical diseases 2021–2030: what we know and what we need to do for global control. Trop Med Health. 2021 Dec;49(1):64.

26. Kabona G. The World Health Organization Road Map for Neglected Tropical Diseases 2021-2030: Will Tanzania achieve the target? | EBSCOhost. Tanzan J Health Res. 2022; 23:213.

27. Ministry of Health. NTD Sustainability plan final 24th Jan 2022 [Internet]. MoH; 2022 [cited 2026 Jan 23]. Available from: https://www.moh.go.tz/storage/app/uploads/public/620/b58/bc2/620b58bc2761a915449308.pdf

28. Ministry of Health. MEDIUM TERM STRATEGIC PLAN (2021/22-2025/26 [Internet]. 2021 [cited 2026 Jan 23]. Available from: https://www.moh.go.tz/storage/app/uploads/public/687/9f6/3fa/6879f63fad8f2963963184.pdf

29. Girma A, Abdu I, Teshome K. Prevalence and determinants of scabies among schoolchildren in Africa: A systematic review and meta-analysis. SAGE Open Med. 2024; 12:20503121241274757.

30. World Health O. Target product profile for scabies to start and stop mass drug administration. 2022;14.

31. Joseph M, Mushi V, Palolo H, Silvestri V, Kinabo C, Mshana I, et al. Prevalence of Sarcoptes scabiei infestation and its associated factors among primary school children: A school-based cross-sectional survey in the Rufiji district, Tanzania. IJID Reg. 2024;11:100365.

32. Dagne H, Dessie A, Destaw B, Yallew WW, Gizaw Z. Prevalence and associated factors of scabies among schoolchildren in Dabat district, northwest Ethiopia, 2018. Environ Health Prev Med. 2019 Dec;24(1):67.

33. Ejigu K, Haji Y, Toma A, Tadesse BT. Factors associated with scabies outbreaks in primary schools in Ethiopia: a case–control study. Res Rep Trop Med. 2019 Aug 27;119–27.

34. Amare HH, Lindtjorn B. Risk factors for scabies, tungiasis, and tinea infections among schoolchildren in southern Ethiopia: A cross-sectional Bayesian multilevel model. PLoS Negl Trop Dis. 2021;15(10):e0009816.

35. Ekuerhare BE, Umukoro EK, Onakpoya EE, Moke EG, Demaki WE, Isibor NP. Prevalence and Knowledge of Scabies among Students Residing in University Hostels of Delta State University, Abraka, Nigeria. Trop J Health Sci. 2024;31(2):20–5.

36. Ugbomoiko US, Oyedeji SA, Babamale OA, Heukelbach J. Scabies in Resource-Poor Communities in Nasarawa State, Nigeria: Epidemiology, Clinical Features and Factors Associated with Infestation. Trop Med Infect Dis. 2018 June 4;3(2):59.

37. Akunzirwe R, Agaba B, Kizito S, Bulage L, Kwesiga B, Migisha R, et al. An outbreak of scabies in a fishing community in Hoima District, Uganda, February−June, 2022. Res Sq Res Sq. 2023;

38. Sara J, Haji Y, Gebretsadik A. Scabies Outbreak Investigation and Risk Factors in East Badewacho District, Southern Ethiopia: Unmatched Case Control Study. Dermatol Res Pract. 2018 June 26;2018:e7276938.

39. Wochebo W, Haji Y, Asnake S. Scabies outbreak investigation and risk factors in Kechabira district, Southern Ethiopia: unmatched case control study. BMC Res Notes. 2019 May 29;12(1):305.

40. Shifera N, Yosef T. Burden and determinants of scabies in a pastoralist community: a case–control study from Southwest Ethiopia. BMJ Open. 2024;14(11):e087097.

41. Mwageni N, Schoenmakers A, van Wijk R, Kamara DV, Kisonga RM, Njako B, et al. Skin diseases, including skin NTDs, identified through community-based integrated skin screening: Findings from the PEP4LEP study in Tanzania. medRxiv. 2025;2025.12. 02.25341087.

42. Melese F, Malede A, Sisay T, Geremew A, Gebrehiwot M, Woretaw L, et al. Cloth sharing with a scabies case considerably explains human scabies among children in a low socioeconomic rural community of Ethiopia. Trop Med Health. 2023;51(1):52.

43. Jira SC, Matlhaba KL, Mphuthi DD. Evaluating the current management approach of scabies at selected primary health care in the Deder district, Ethiopia. Heliyon. 2023;9(1).

44. Gezmu T, Enbiale W, Asnakew M, Bekele A, Beresaw G, Nigussie M, et al. Does training of Health Extension Workers reduce scabies load in district health facilities in rural Ethiopia? J Infect Dev Ctries. 2020 June;14(06.1):36S–41S.

45. Padovese V, Dassoni F, Morrone A. Scabies coexisting with other dermatoses: the importance of recognizing multiple pathologies in resource-poor settings. Int J Dermatol. 2020;59(12):1502–5.

46. Azene AG, Aragaw AM, Wassie GT. Prevalence and associated factors of scabies in Ethiopia: systematic review and Meta-analysis. BMC Infect Dis. 2020 Dec;20(1):380.

