## Supplemental 1 and will be used for the link to the file on the preprint site. for "Behavioral and Healthcare Determinants of Self-Reported Scabies in Chiwanda Ward, Nyasa District, Tanzania"

### THE UNITED REPUBLIC OF TANZANIA

National Institute for Medical Research
3 Barack Obama Drive
P.O. Box 9653
11101 Dar es Salaam


Permanent Secretary
Ministry of Health
Government City Mtumba
Health Road
P.O. Box 743
40478 Dodoma

NIMR/HQ/R.8a/Vol.IX/4583

04 April 2024

Mr. Ibrahim Twahir Kilagwa
Sokoine University of Agriculture
College of Veterinary Medicine and Biomedical Sciences
P O Box 3021
Morogoro

**RE: ETHICAL CLEARANCE CERTIFICATE FOR CONDUCTING MEDICAL RESEARCH IN TANZANIA**

This is to certify that the research entitled: **"Leveraging AfyaData Digital Technology for Scabies Outbreak Surveillance in Nyasa District, 2022" (Kilagwa, I. *et al*)** has been granted ethical clearance to be conducted in Tanzania.

The Principal Investigator of the study must ensure that the following conditions are fulfilled:

1. Progress reports are submitted to the Ministry of Health and the National Institute for Medical Research, Regional and District Medical Officers after every six months.
2. Permission to publish the results is obtained from the National Institute for Medical Research.
3. Copies of final publications are made available to the Ministry of Health and the National Institute for Medical Research.
4. Any researcher, who contravenes or fails to comply with these conditions, shall be guilty of an offence and shall be liable on conviction to a fine as per NIMR Act No. 23 of 1979, PART III Section 10(2).
5. Sites: Ruvuma region.

Approval is valid for one year from 04 April 2024 to 03 April 2025.

**Name: Prof. Said S. Aboud**

**Signature**
**CHAIRPERSON**
**MEDICAL RESEARCH**
**COORDINATING COMMITTEE**

**Name: Prof. Tumaini J. Nagu**

**Signature**
**CHIEF MEDICAL OFFICER**
**MINISTRY OF HEALTH**

c.c: Director, Health Services-TAMISEMI, Dodoma.
RMO of Ruvuma region.
DMO/DED of Nyasa district.
