## Supplemental 2, and will be used for the link to the file on the preprint site. for "Behavioral and Healthcare Determinants of Self-Reported Scabies in Chiwanda Ward, Nyasa District, Tanzania"

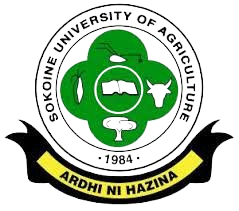

**SECTION 1: DEMOGRAPHICS**

**This section will constitute information about you, cross [X] appropriately.**

1. Household GPRS Coordinates. ______________________________________
2. What is the village you represent? _______________________________________
3. What is your Gender: □ Male □Female
4. What is your level of education?□ No education□ Primary □High school □College□ University
5. Do you have a job:□YES□ NO

If Yes Specify ……………………………………………………………………………….

6. Religious affiliation:□ Protestant □Muslim □ Roman Catholic□ SDA□ African Tradition Religion
7. MaritalStatus:□MarriedMonogamy □Married polygamy□Widow/Widower□Divorce/Separated

**SECTION 3: SCABIES OUTBREAK TIMELINE**

8. Where you aware of the scabies outbreak in your village ?□YES□ NO
9. When did you first become aware of the scabies outbreak in 2022, Nyasa? ……………………………......................................................................................................
10. Did you or your household experience symptoms of scabies during the outbreak?□YES□ NO

If yes, please describe the regions of the body where the itching and rash were most severe.

□ Wrist□ Elbow □ armpits □ genitals □ buttocks□ fingers □ toes

**SECTION 4: ENVIRONMENTAL FACTORS**

11. Could you please provide the number of individuals currently living in your household

□ under 5 □ above 5

12. How often do you shower a day?□ Once□ Twice □ Three times or more
13. What is the source of water used for showering □ Rain □protected well□ un-protected well □ Lake □ RUWASA □ Spring
14. Do you keep any Animals in your household □YES□ NO

If yes, what types of animals do you have in your household □Dog□ Cat □Pig□ Cow

15. How often do you bathe your animal/ pet? □Once a week □ a few times months □rarely
16. Have you or your household members been in close contact with animals during the outbreak? □YES□ NO

If yes, please specify the types of animals and the extent of interaction

…………………………………………………………………………………………………

…………………………………………………………………………………………………

**SECTION 5: INTERVENTION AND TREATMENT**

17. Was there any specific treatment or medication provided during the scabies outbreak

□YES□ NO □Not sure

If yes, please specify the names of the medications if known:

□Oral medication □ Topical creams

18. Were any preventive measures communicated or implemented to control of scabies □YES□NO

If yes, please specify the preventive measures taken:

□Education on personal hygiene □ Isolation of affected individuals □Cleaning and disinfecting living spaces

19. Was there collaboration with healthcare professionals or organizations during the outbreak □YES□ NO □Not sure Environmental Factors-Environmental- Environmental
