## Supplemental Table 3, and will be used for the link to the file on the preprint site. for "Behavioral and Healthcare Determinants of Self-Reported Scabies in Chiwanda Ward, Nyasa District, Tanzania"

**SCABIES CASE DEFINITION AND ALERT CRITERIA FOR HOUSEHOLD SURVEILLANCE**

*Community-based screening, alert triggering, and reporting guidance*

### 1. Purpose

To provide a clear, practical case definition for identifying suspected scabies during household visits, raising an alert in the community, and aligning household surveillance with standard clinical and epidemiological reporting concepts.

### 2. Household Screening Signs (for community household visits)

Use the following three screening questions/signs for each assessed household member:

a) Visible rash on the skin (Yes/No)
b) Itching that is worse at night / Severe nocturnal itching (Yes/No)
c) One or more household members have similar symptoms (Yes/No).

### 3. Household Alert Case Definition (community alert trigger)

A household should raise an alert when any one assessed household member meets all three screening criteria (rash + nocturnal itching + household member(s) with similar symptoms).

**Alert checklist (tick Yes/No for the assessed person):**

| **Criterion** | **Response (Yes/No)** |
| --- | --- |
| Criterion 1: Visible rash | YES |
| Criterion 2: Itching worse at night | YES |
| Criterion 3: One or more household members with similar symptoms | YES |

**Alert Trigger: Trigger an alert if all three criteria are answered “Yes” for any one assessed household member.**

### STANDARD CASE DEFINITIONS (CLINICAL & SURVEILLANCE REPORTING)

#### Suspected scabies case

A person with pruritus (itching), especially worse at night, with characteristic skin lesions (e.g., papules, vesicles, excoriations, or burrow-like lesions), with or without a history of close contact with a scabies case.

#### Confirmed scabies case

A person in whom scabies mites, eggs, or mite feces are identified by microscopic examination of skin scrapings or by visualization of mites through dermoscopy.

### 4. Epidemiological Criteria (individuals at risk)

- Close contact with a suspected or confirmed scabies case (especially within the same household).
- Residence in, or recent stay in, crowded or institutional settings (e.g., dormitories, prisons, camps, boarding schools).

### 5. Common Lesion Sites (to guide examination)

- Interdigital spaces, Wrist, Elbow, Axilla, Beltline/waist, Genital area, Buttocks

### 6. Case Classification (during investigations)

- **Index case:** The first identified case in a group, household, or community.
- **Secondary case**: A person with scabies who is epidemiologically linked to the index case.

### 7. Exclusion Criteria

Exclude cases where another diagnosis clearly explains the symptoms and skin lesions.

### 8. Surveillance and Reporting

Health facilities and community surveillance teams should promptly report suspected or confirmed scabies cases to the relevant local health authorities according to routine surveillance procedures.

### 9. Control Measures (basic)

- Treat affected individuals according to national/clinical guidance.
- Treat close contacts (especially household members) to reduce re-infestation and ongoing transmission.
- Provide health education on personal hygiene, environmental cleaning, and correct use of medications.
- Encourage early care-seeking for symptomatic individuals.
