## Supplemental 5, and will be used for the link to the file on the preprint site. for "Behavioral and Healthcare Determinants of Self-Reported Scabies in Chiwanda Ward, Nyasa District, Tanzania"

SOKOINE UNIVERSITY OF AGRICULTURE (SUA)

SACIDS Africa Centre of Excellence for Infectious Diseases of
Humans and Animals in Eastern and Southern Africa
College of Veterinary Medicine and Biomedical Sciences
P.O Box 3015, Chuo Kikuu, Morogoro, Tanzania

url: www.sacids.org

Our ref: SACIDS/ADM/23/237 | Your ref: | 10th November, 2023

To

Whom it may concern

**REF: Support letter for Mr. Kilagwa Ibrahim**

I hereby grant approval for the use of the *AfyaData* app for the MSc research under the following conditions:

1. Ethical Procedures: It is imperative that the research strictly adheres to all ethical guidelines and procedures applicable to data collection, ensuring the highest standards of integrity and compliance.

2. Acknowledgment: I kindly request that Mr. Kilagwa and his research team acknowledge the SACIDS Foundation for One Health as the rightful owner of the *AfyaData* tool in all future research reports, publications, and any dissemination of research findings. Proper attribution is vital to recognizing our organization's contributions to this research endeavor.

Should you require any further clarification or have questions regarding this authorization or related matters, please do not hesitate to contact me directly.

I have full confidence that this authorization will facilitate the successful execution of MSc research project and contribute significantly to the field of health data science.

Thank you for your cooperation and commitment to ethical research practices.

SACIDS FOUNDATION FOR ONE HEALTH
ONE AFRICA, ONE HEALTH
P. O. BOX 3297
MOROGORO

**Neema Heri Mwakabonga**
Administrator Cum Executive Assistant
SACIDS Foundation for One Health

CamScanner
